# Genetic risk of late-onset Alzheimer’s disease is associated with longitudinal loss of functional brain network segregation in middle-aged cognitively healthy individuals: The PREVENT-Dementia Study

**DOI:** 10.1101/2023.04.18.23288690

**Authors:** Feng Deng, Karen Ritchie, Graciela Muniz-Terrera, Paresh Malhotra, Craig W. Ritchie, Brian Lawlor, Lorina Naci

## Abstract

It is well acknowledged that the pathological processes of Alzheimer’s disease (AD) start decades before clinical manifestations, but early indicators of AD in midlife remain unclear. Functional segregation of brain networks has recently emerged as a key indicator of brain health. In this study, we investigated the vulnerability of intrinsic brain networks to loss of functional segregation during healthy adult lifespan and in cognitively healthy midlife individuals at risk of late-onset AD, and the association between segregation loss and cognition in midlife. Network segregation was measured using the participation coefficient metric within a graph-theoretic framework. In a healthy adult lifespan cohort (18-88 years, N=652), linear relationships of network segregation with age and cortical grey matter volume (GMV) were assessed using multiple regression models. In a cognitively healthy midlife cohort (40-59 years, N=210), associations between network segregation and established risk factors for AD were examined cross-sectionally and longitudinally (over 2 years). Across the healthy adult lifespan, global network segregation was positively associated with GMV and negatively associated with age, replicating previous findings. Three high-order networks [default mode (DMN), frontal-parietal control, and salience] and two sensorimotor networks (visual and motor) showed prominent age-related changes in functional segregation throughout adulthood. At midlife, cross-sectionally, cognitively healthy apolipoprotein (*APOE*) ε4 carriers had higher global segregation than non-carriers. The DMN was the only individual network to show such an effect of *APOE* genotype. Higher global and DMN segregation was associated with better episodic and relational memory. Critically, *APOE* ε4 carriers, but not non-carriers, showed a significant longitudinal loss of segregation in the DMN over 2 years. Overall, our findings suggest that functional network segregation constitutes a novel and early substrate for the impact of the genetic AD risk on the brain in midlife and thus have implications for the early detection and intervention in AD.

## 1 Introduction

Dementia, particularly Alzheimer’s disease (AD), is a growing public health issue that presents profound challenges to healthcare systems, families, and societies throughout the world (World Health Organization, 2021). Midlife is a critical period for the development of AD pathology (Jansen et al., 2015; Sperling et al., 2011) and potentially a unique disease-altering window prior to the manifestation of substantial brain damage. Therefore, there is an urgent need for risk reduction interventions focused on midlife (Barnes & Yaffe, 2011; Livingston et al., 2017; Ritchie et al., 2010). However, the indicators and brain mechanisms of AD in midlife remain poorly understood (Irwin et al., 2018; Ritchie et al., 2017).

The brain is composed of intrinsically wired functional networks (Crossley et al., 2013; Smith et al., 2009), each corresponding to a set of distinct and tightly connected regions (Cole et al., 2014; Smith et al., 2009), often involved in specialised functional processing (Sporns & Betzel, 2016; Wig, 2017). Such modular functional organisation of the brain in the form of distinct networks is critical for cognition (Achard et al., 2006; Bullmore & Sporns, 2012; Chan et al., 2014). For example, across the adult lifespan, individuals with greater segregation of functional brain networks show better long-term episodic memory (Chan et al., 2014). Therefore, disrupted network segregation may lead to cognitive and behavioural decline. There are several factors and/or conditions associated with disruption of functional network segregation and that have implications for cognition. In healthy populations, increasing adult age is associated with reduced network segregation or more diffuse functional organisation of the brain (Chan et al., 2014; Wig, 2017). Such ‘dedifferentiation’ of functional networks is in turn associated with age-related decline in cognitive and motor function (Chan et al., 2014; King et al., 2018; Kong et al., 2020; Manza et al., 2020; Pedersen et al., 2021; Varangis et al., 2019). Conversely, preservation of network segregation is associated with maintenance of cognition in healthy ageing (Cassady et al., 2020; Chan et al., 2021; Gallen et al., 2016) and in patients with brain injury (Arnemann et al., 2015), suggesting that functional network segregation supports cognitive reserve (Stern, 2012).

In asymptomatic older adults (mean age > 65 years), loss of network segregation is associated with the accumulation of AD pathology, such as beta-amyloid (Aβ) and tau (Brier et al., 2014; Ewers et al., 2021), or the presence of the Apolipoprotein E (*APOE*) ε4 allele (Ng et al., 2018), the main genetic risk factor for sporadic late-onset AD in the Indo-European population (Lambert et al., 2013), compared to the absence of these conditions. These findings suggest that network segregation is impaired in cognitively unimpaired older adults at risk for AD. This reduction in network segregation in the high-risk group was further associated with cognitive decline (Ng et al., 2018), whereas maintenance of network segregation was associated with preserved cognitive performance, despite the presence of AD pathology (Ewers et al., 2021). Furthermore, decreases in network segregation have been observed in patients with MCI (Farràs-Permanyer et al., 2019; Jiao et al., 2021) and AD (Dai et al., 2019; Ewers et al., 2021) compared to age-matched controls. Importantly, an accelerated decline in network segregation with ageing was associated with increasing dementia severity (Chan et al., 2021). Taken together, the accumulating evidence points to network segregation as a marker of brain health in both normal and pathological ageing.

An important question that remains to be addressed is the selective vulnerability of individual networks to loss of functional segregation. Studies suggest that large-scale brain systems undergo nonuniform changes during healthy ageing, with the associative system being more susceptible to age-related ‘dedifferentiation’ than the sensorimotor system (Betzel et al., 2014; Chan et al., 2014; Geerligs et al., 2015; Pedersen et al., 2021; Siman-Tov et al., 2017; Wig, 2017). However, it remains unclear which individual networks within the associative system are more liable to segregation loss during healthy ageing and in preclinical AD populations.

Furthermore, it remains unknown whether the risk of late-onset AD is associated with altered functional network segregation in cognitively healthy, middle-aged individuals, who may be decades before clinical manifestations. Answers to this question have important implications for identifying intermediate phenotypes of the earliest brain changes in the preclinical stages of AD (Foo et al., 2020), which will help to provide urgently needed early disease biomarkers in the earliest stages of the disease, and complement previous studies summarised above to elucidate the earliest time point for network segregation changes along the AD spectrum.

To address these two research gaps, the first aim of this study was to investigate age-related differences in functional segregation of individual networks in a large healthy adult lifespan cohort from the Cambridge Centre for Ageing and Neuroscience (Cam-CAN) open dataset (N = 652, 18-88 years). The second aim was to test hypotheses derived from the results of the healthy ageing investigation, to examine the impact of three risk factors for late-onset Alzheimer’s disease, i.e., *APOE* ε4 allele, family history of dementia (FHD) (Scarabino et al., 2016) and Cardiovascular Risk Factors Aging and Dementia (CAIDE) score (Kivipelto et al., 2006), on network segregation in midlife. Cross-sectional and longitudinal (over 2 years) changes in network segregation and their associations with cognition were examined in a midlife cohort of cognitively healthy individuals from the PREVENT-Dementia research programme (N = 210, 40-59 years).

Our first prediction was that there would be differential age effects on ten predefined brain networks based on a comprehensive whole-brain atlas (Power et al., 2011) across the healthy adult lifespan (the Cam-CAN cohort), with high-order networks in particular expected to show the strongest age effect. The second prediction was that in the midlife PREVENT-Dementia cohort, the global segregation of functional networks, and the segregation of networks most susceptible to the healthy ageing processes would be influenced by the risk of late-onset AD. Furthermore, studies have shown that the DMN is particularly vulnerable to AD pathology (Greicius et al., 2004; Kucikova et al., 2021; Márquez & Yassa, 2019; Rombouts et al., 2005). For example, atrophy and metabolic abnormalities occur in the core regions of the DMN at early stages of AD progression (Buckner et al., 2005; Dickerson et al., 2009; Minoshima et al., 1997). Therefore, we expected that functional segregation of the DMN in particular would be affected by AD risk in the middle-aged cohort. Specifically, we tested three hypotheses in the midlife cohort: (i) reduced global network segregation in the high-risk groups compared to the low-risk group cross-sectionally, (ii) a greater reduction in high-order networks, particularly in the DMN and (iii) a more pronounced longitudinal decline over 2 years in the high-risk group compared to the low-risk group.

## 2 Methods

### 2.1 Participants

#### Healthy lifespan adults

The healthy lifespan cohort was drawn from the Cam-CAN research programme (http://www.cam-can.org/), a large-scale collaborative research project aimed at elucidating the neurocognitive mechanisms that underpin healthy cognitive ageing. A detailed protocol has been described elsewhere (Shafto et al., 2014). Ethical approval was obtained from the Cambridgeshire 2 (now East of England-Cambridge Central) Research Ethics Committee, and informed consent was obtained from all participants prior to assessments and imaging. The cohort consisted of healthy volunteers aged 18-88 years. Data collected in the second phase of this project were examined in the present study. 652 participants (322 male; 330 female) underwent structural and resting-state fMRI scans. All participants have fMRI data and 646/652 have structural MRI data after quality control.

#### Middle-aged adults at risk for late-onset AD

The cognitively healthy middle-aged cohort was recruited from the PREVENT-Dementia research programme, an ongoing longitudinal multi-site research programme across the UK and Ireland, seeking to identify early biomarkers of AD and elaborate on risk-mechanism interactions for neurodegenerative diseases decades before the cardinal symptoms of dementia emerge. Its protocol has been described in detail elsewhere (Ritchie & Ritchie, 2012). In the first PREVENT programme phase, participants were recruited at a single site, via the dementia register database held at the West London National Health Service (NHS) Trust, of the UK National Health Service, the Join Dementia Research website (https://www.joindementiaresearch.nihr.ac.uk/), through public presentations, social media and word of mouth. Procedures involving experiments on human subjects were carried out in accord with the ethical standards of the Institutional Review Board of Imperial College London and in accord with the Helsinki Declaration of 1975. Approval for the study was granted by the NHS Research Ethics Committee London Camberwell St Giles. Consented participants were seen at the West London NHS Trust, where they underwent a range of clinical and cognitive assessments (Ritchie & Ritchie, 2012). The cohort comprised cognitively healthy volunteers aged 40-59 years. Here we examined baseline and follow-up data from the West London dataset. 210 individuals (62 male; 148 female) were tested at baseline, with 188 (89.5%) (55 male; 133 female) retained at 2 years follow-up.

At baseline, 17 participants were excluded due to lack of participation or contraindications to MRI, 6 due to incidental findings, and 20 due to inadequate brain coverage (for details please see the following section on the *functional brain network construction*). At follow-up, 19 participants were excluded due to decline or contraindications to MRI, 3 due to incidental findings, and 1 due to inadequate brain coverage. In addition, 21 participants were further excluded from the longitudinal analyses, due to missing either the baseline or the follow-up sessions. Therefore, the dataset for the cross-sectional analyses (i.e., baseline session) was N = 167, and the dataset for the longitudinal analyses (i.e., remaining at both baseline and follow-up sessions) was N = 144 (Figure 1).

**Figure 1.**
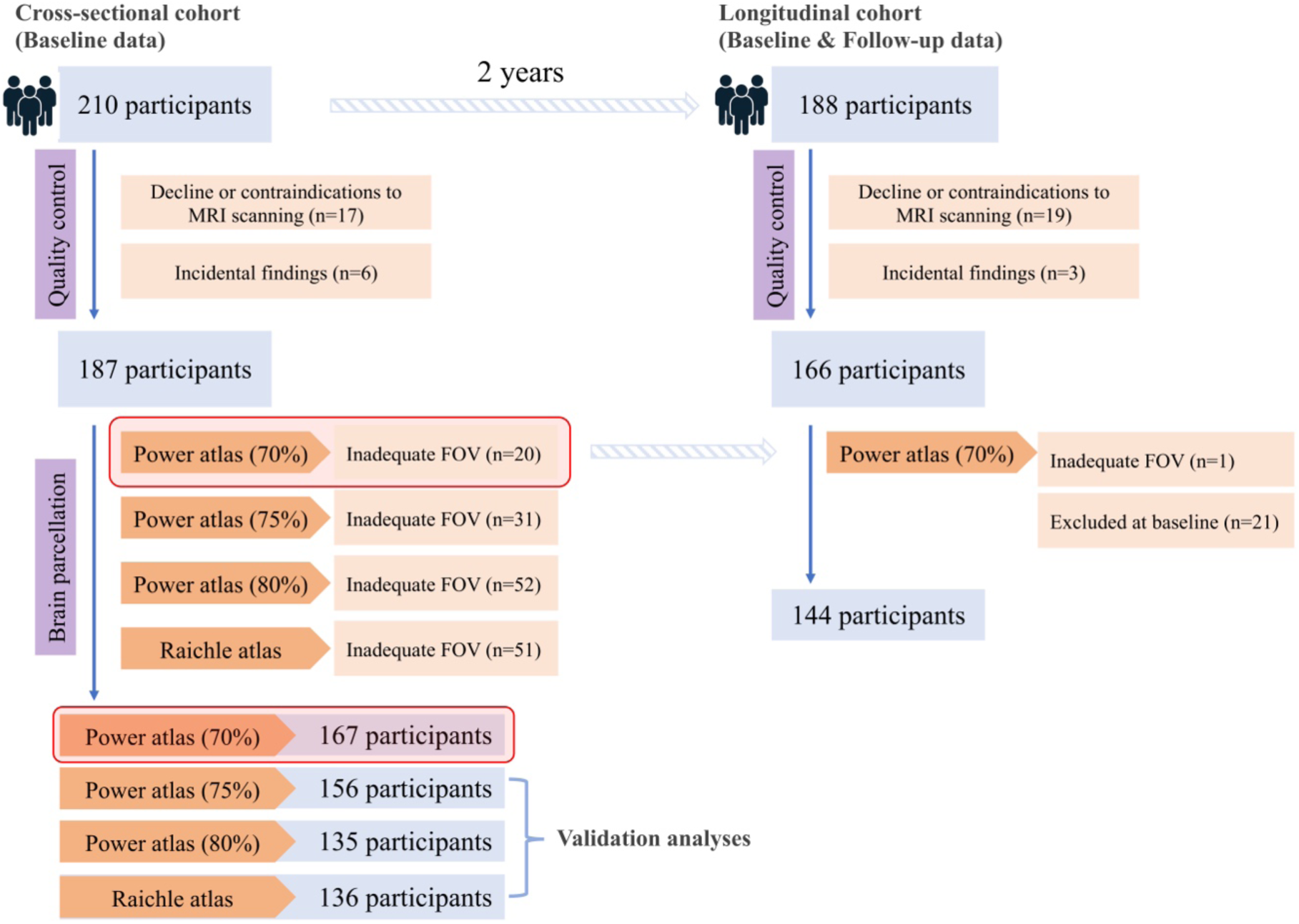
Participant exclusions for resting-state fMRI data analyses of the PREVENT-Dementia cohort. Two brain parcellation schemes were adopted to define the nodes of the brain networks: Power atlas (Power et al., 2011) and Raichle atlas (Raichle, 2011). Due to inadequate field of view (FOV) for Power atlas, participants with less than 70% of the initial brain nodes (node = 214) were excluded. To test if the results were driven by a particular subset of participants, we also tested two more stringent thresholds: 75% and 80%. Due to inadequate FOV for Raichle atlas, participants without full coverage of the whole brain nodes (node = 33) were excluded.

### 2.2 Assessments from the PREVENT-Dementia study

Three risk factors for late-onset AD (*APOE* ε4 allele, FHD, and CAIDE score) were assessed in this study. The details are described in Deng et al. (2022); Ritchie et al. (2017). Briefly, *APOE* ε4 risk is determined by having ≧1 *APOE* ε4 allele; FHD risk is determined by having at least one parent diagnosed with dementia; CAIDE is a composite scale of estimated future dementia risk based on midlife cardiovascular measures (Fayosse et al., 2020; Sindi et al., 2015). It takes into account an individual’s age, sex, educational attainment, *APOE* ε4 genotype, activity level, body mass index, cholesterol and systolic blood pressure (Kivipelto et al., 2006) and is scored on a range of 0-18. A higher score indicates a higher risk. The CAIDE dementia risk score was calculated at baseline and follow-up.

Cognitive function was assessed at baseline and follow-up using the COGNITO neuropsychological battery (Ritchie, 2014), which is designed to examine information processing across a wide range of cognitive functions in adults of all ages and is not restricted to those functions usually implicated in dementia detection in older adults. Additionally, we used the Visual Short-Term Memory Binding Task (VSTMBT), which is sensitive to detecting changes in the pre-symptomatic stages of AD (Parra et al., 2010). In total, 13 measures were derived to capture multiple cognitive functions [see Supplementary Information (SI) for details]. In an independent study of this dataset (Deng et al., 2022), a dimensionality reduction method, i.e., rotated principal component analysis (rPCA), was adopted to cluster these measures into three cognitive components (Supplementary Figure 1) in order to reduce the number of multiple comparisons between the cognitive measures (Jolliffe & Cadima, 2016). This step maximized the statistical power to examine the brain– behaviour relationships. The rPCA was conducted using the psych package (version 2.0.12) in R software (https://www.r-project.org/) and included the following steps: (a) component estimation by using scree plots and parallel analysis, (b) component extraction by using principal component analysis, (c) Varimax rotation to constrain the components to be uncorrelated, and (d) calculation of component scores by a regression method (see Deng et al. (2022) for details). In subsequent analyses, we used the three cognitive components, namely (i) episodic and relational memory, (ii) working and short-term (single-feature) memory, and (iii) verbal and visuospatial functions, and short-term (conjunctive) memory.

### 2.3 MRI data acquisition and pre-processing

#### The Cam-CAN study

Imaging data were collected at a single site (MRI-CBSU) using a 3T Siemens TIM Trio scanner with a 32-channel head coil. Full descriptions of the MRI protocols have been described elsewhere (Taylor et al., 2017). Resting state fMRI data were acquired using a T2*-weighted echo planar imaging (EPI) sequence with participants resting with their eyes closed. 261 volumes were acquired, and each volume contained 32 axial slices (in descending order) with a slice thickness of 3.7 mm and an interslice gap of 20% [repetition time (TR) = 1970 ms, echo time (TE) = 30 ms, flip angle (FA) = 78°, field of view (FOV) = 192 × 192 mm^2^, voxel size = 3 mm × 3 mm × 4.44 mm]. A 3D T1-weighted magnetization prepared rapid gradient-echo image (MPRAGE, TR = 2250 ms, TE = 2.99 ms, FA = 9°, FOV = 256mm × 240mm × 192mm, voxel size = 1 mm^3^ isotropic) was also acquired.

Resting-state fMRI data were preprocessed by the Cam-CAN group using a standard preprocessing pipeline with statistical parametric mapping (SPM12, https://www.fil.ion.ucl.ac.uk/spm/software/spm12/) and automatic analysis (AA) software (Cusack et al., 2014). Details of the pipeline are described in Taylor et al. (2017). Briefly, raw fMRI data were unwarped using field map images for distortion correction due to magnetic field inhomogeneities, realigned for motion correction, and corrected for slice timing. Functional images were then coregistered with T1 structural images and normalised to Montreal Neurological Institute (MNI) standard space using normalisation parameters derived from the Diffeomorphic Anatomical Registration through Exponentiated Lie Algebra (DARTEL) procedure (Ashburner, 2007). To further account for the effects of head motion, a wavelet despiking method was applied to remove motion artefacts (Patel et al., 2014).

In addition, we applied a general linear model (GLM) including 24 head motion parameters and white matter (WM) and cerebrospinal fluid (CSF) signals to reduce residual effects of head motion and other noise confounders. The 24 parameters included six original rigid-body motion parameters, the first-order temporal derivatives of these six parameters, and 12 quadratic terms of the original motion parameters and their derivatives (Satterthwaite et al., 2013). Frame-wise displacement (FD) motion parameters (Power et al., 2012) were calculated as the sum of the absolute values of the differentiated realignment estimates at each time point (see SI for full descriptions and the formula), which measures the movement of the head from one volume to the next (Power et al., 2012). We then averaged the FD across time points and regressed it in the group-level analyses to further account for head movement. Finally, high-pass temporal filtering (Gaussian-weighted least-squares straight line fitting, corresponding to 100 s) was applied to remove low-frequency artefacts. Spatial smoothing was not applied for network analysis, as suggested by Alakörkkö et al. (2017).

Morphometric brain measures were derived from the T1 images using the Mindboggle pipeline (Klein et al., 2017). Grey matter volume (GMV) of 34 brain regions per hemisphere based on the Desikan-Killiany atlas (Desikan et al., 2006) was extracted using the underlying Freesurfer processing pipeline (Klein & Tourville, 2012). The mean GMV of these cortical areas was used in this study to represent the structural integrity.

#### The PREVENT-Dementia study

Imaging data were obtained as part of a multimodal examinations in a 3T Siemens Verio MRI scanner and with 32-channel head coil (https://preventdementia.co.uk/for-researchers/). Resting-state fMRI data were acquired with T2*-weighted EPI sequence. 330 volumes were acquired, and each volume contained 35 slices (interleaved acquisition), with slice thickness of 3 mm (TR = 2000ms, TE = 30ms, FA = 80°, FOV = 192 × 192mm^2^, voxel size = 3 mm^3^ isotropic). A 3D T1-weighted MPRAGE image (160 slices, voxel size = 1 mm^3^ isotropic, TR = 2300ms, TE = 2.98ms, FOV = 240 × 256mm^2^, FA = 9°) was also acquired. All scans were repeated after approximately 2 years on the same scanner using the same protocol.

Standard preprocessing procedures for resting-state fMRI data were performed with SPM12 and AA software (Cusack et al., 2014) implemented in MATLAB R2019a (The MathWorks, United States). In this pipeline (Figure 2a), we performed slice timing correction, motion correction, co-registration of functional and structural images, normalization into standard MNI space, and spatial smoothing. Spatial normalization was performed using SPM12’s segment-and-normalize procedure, whereby the T1 structural was segmented into GM, WM and CSF and normalized to a segmented MNI-152 template. These normalization parameters were then applied to all EPIs. The quality of spatial normalization was visually inspected for each participant and no participants showed a normalization failure. The data were then smoothed with a Gaussian kernel of 6mm full width at half maximum and were temporally band-pass filtered (0.01-0.08 Hz) to remove low-frequency drift and high-frequency physiological noise (Salvador et al., 2008; Zuo et al., 2010). Finally, to reduce any residual effects of head movement, a GLM was applied with the 24 head movement parameters mentioned above included as covariates (Satterthwaite et al., 2013). In addition, mean FD was calculated and regressed in the group level analyses (Power et al., 2012).

**Figure 2.**
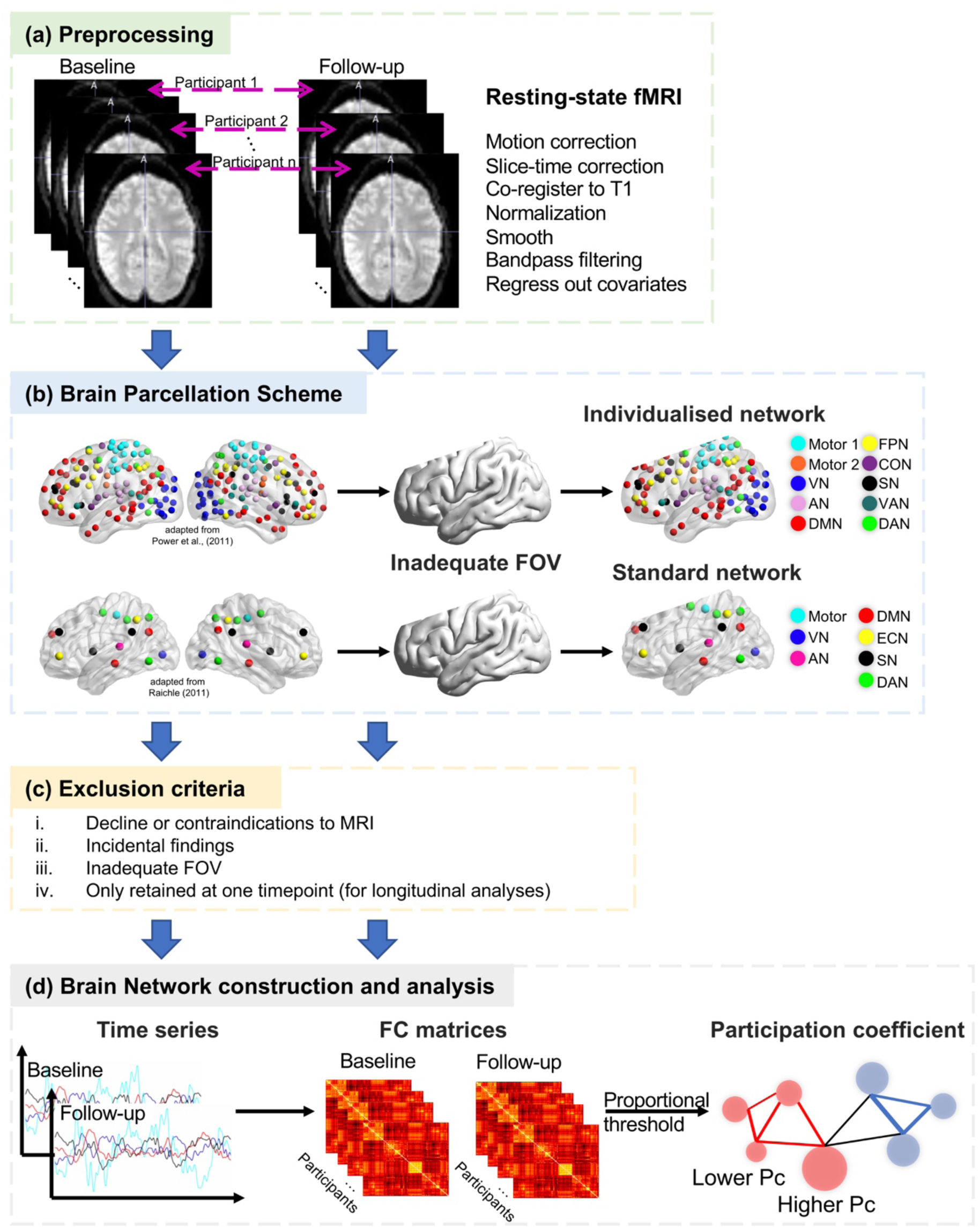
The schematic of the study design for the PREVENT-Dementia study. Resting state fMRI data were collected from a cognitively healthy middle-aged cohort at baseline (N = 210; aged 40-59 years old), and some of them (N = 188) were followed up over 2 years. (a) Standard preprocessing steps were performed separately for the baseline and follow-up datasets. (b) Two brain parcellation schemes were adopted to define the nodes of the brain networks. Due to the inadequate field of view (FOV) and different scanning angles that were set up for each participant’s fMRI scan, individualised brain networks were adapted from a comprehensive brain atlas (Power et al., 2011), comprising 10 predefined networks, according to the coverage of individual-specific functional brain images. In addition, we adopted another brain parcellation scheme (Raichle, 2011) that comprises a smaller number of key regions for 7 predefined networks to validate the main results. In this atlas, we can ensure the same set of brain nodes for every participant. (c) Several criteria were applied to exclude the inappropriate data for the network analyses. (d) Time courses were extracted based on the brain nodes in the individual-specific atlas and correlated with each other to create functional connectivity (FC) matrices by using Pearson correlation (r). Proportional thresholds were applied to FC matrix to generate the sparse matrix for the calculation of participation coefficient (Pc) to describe functional segregation. The mock graph illustrates two different networks in red and blue. The node (in red) with lower Pc exhibits strong connections only within its belonging network (edges in red), but no connection to the other network (in blue), indicating higher functional segregation. By contrast, the node (in red) with higher Pc exhibits equally distributed connections to its belonging network (edges in red) and to the other network (edges in black), indicating lower functional segregation or more diffused brain. Abbreviations: VN, visual; AN, auditory; DMN, default mode; FPN, frontal-parietal control; SN, salience; VAN, ventral attention; DAN, dorsal attention; CON, cingulo-opercular control; ECN, executive control.

### 2.4 Resting-state fMRI data analyses

#### Functional brain network construction

A graph-theoretic framework was adopted to guide analyses of the functional organisation of brain networks using resting-state fMRI data.

##### Node definition

214 brain nodes (spherical, 5 mm diameter) were defined based on a previously published functional system map (Power et al., 2011) comprising 10 functional brain networks: motor 1 (sensorimotor hand), motor 2 (sensorimotor mouth), visual, auditory, default mode (DMN), frontal-parietal control (FPN), cingulo-opercular control (CON), ventral attention (VAN), dorsal attention (DAN) and salience network (SN).

The resting-state fMRI data from the PREVENT cohort have an insufficient field of view (FOV) to cover the whole brain, resulting in the exclusion of some brain nodes in this functional brain map (Power et al., 2011). In addition, different fMRI scan angles were set for different participants, resulting in different FOVs that prevented exclusion of the same set of brain nodes for each participant. To overcome these limitations, we excluded different sets of brain nodes from this brain map based on participants’ specific FOV and constructed individualised brain networks (Figure 2b, see also SI for details).

To ensure that participants with poor brain coverage, leading to a small number of retained brain nodes, did not bias the network analyses, we excluded participants with less than 70% of the original 214 brain nodes. This threshold was chosen to retain a relatively good number of brain nodes and a satisfactory number of participants to ensure statistical power. The number of retained brain nodes was also included as a covariate in the statistical models to further account for its effect.

##### Edge definition

Functional connectivity (FC) between pairs of predefined brain nodes was obtained by calculating the Pearson correlation coefficient *r* of the denoised fMRI time courses derived from these nodes (van den Heuvel & Hulshoff Pol, 2010), forming the FC matrix (Figure 2d). To avoid the formation of artificial anticorrelations, we did not perform a global signal regression (Anderson et al., 2011; Murphy et al., 2009). Negative connectivity was removed due to its ambiguous meaning (Chai et al., 2012; Murphy et al., 2009).

Thresholding the FC matrix to form a sparse matrix is important to remove spurious connections (van den Heuvel et al., 2017). FC matrices were therefore thresholded from 5% to 50% connection density with a 5% interval, as graphs become more random above a threshold of 50% (Humphries et al., 2006). The area under the curve (AUC) for the graph theoretical measure across all thresholds was calculated to provide a scalar that does not depend on a specific threshold selection (Achard & Bullmore, 2007; Wang et al., 2009).

#### Participation coefficient

The participation coefficient (Pc) of a brain node represents the distribution of its connections across separate networks (Guimerà & Nunes Amaral, 2005; Power et al., 2013) and was therefore used to measure the segregation property of the functional brain network. The equation is as follows:

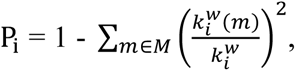

Where *m* is a network in a set of networks *M. k^w^_i_(m)* is the weighted connections of node *i* with all nodes in the network *m. k^w^_i_(m)* is the total weighted connections node *i* exhibits.

P_i_ close to 0 indicates that node *i* is highly segregated, with most of its connections restricted to its own network and a relatively sparse connections to other networks (Figure 2d). In contrast, P_i_ close to 1 indicates that node *i* is highly integrated with nodes of other networks, represented by more equally distributed connections among different networks (Figure 2d). To quantify the segregation of individual networks, we averaged P_i_ across brain nodes that were assigned to the same network. The average P_i_ of nodes across the whole brain was used to quantify global network segregation.

### 2.5 Statistical analyses

#### The Cam-CAN study

All statistical analyses were performed using R software. The demographics of the participants are summarised in Table 1. We used multiple linear regression models to examine the relationship between global network segregation and mean cortical GMV, including age, sex, educational level and mean FD as covariates. The ratio of mean cortical GMV to intracranial volume was calculated for each participant to account for inter-individual differences in head size. To investigate the effect of age on global and individual functional network segregation, global and Pc of each of the ten networks were treated as dependent variables in separate models, with age as the independent variable and sex, education attainment and mean FD as covariates. Multiple comparisons between the 10 individual networks were Bonferroni-corrected.

**Table 1.** Demographic information for the Cam-CAN cohort

|  |  | Whole cohort |
| --- | --- | --- |
| Age range, y |  | 18-88 |
| Age mean ± SD, y | | 54.4 ± 18.5 |
| Sex (Female), % |  | 50.61 |
| Education <sup>a</sup> | University | 395 |
|  | A Levels | 68 |
|  | GCSE | 111 |
|  | None | 75 |

<sup>a</sup> Education levels were categorized according to the British education system: ‘none’, no education over the age of 16 years; ‘GCSE’, General Certificate of Secondary Education; ‘A Levels’, General Certificate of Education Advanced Level; ‘University’, undergraduate or graduate degree. Educational information were missing for 3 participants. SD = standard deviation.

#### The PREVENT-Dementia study

The normality of the data was assessed by combining the visualization of a quantile-quantile plot and the Shapiro-Wilk test. Demographic and clinical information of this study cohort was analysed across risk groups, using 9, tests for categorical variables and Mann-Whitney U tests for continuous variables, given that they were not normally distributed in this cohort (Table 2).

**Table 2.** Demographic information for the PREVENT-Dementia cohort cross-sectionally (at baseline) and longitudinally (over 2 years), stratified by family history of dementia and APOE genotype.

| | | Family history | | | APOE ε4 (2 missing data) | | |
| --- | --- | --- | --- | --- | --- | --- | --- |
| | | FHD+<br>(N=79) | FHD-<br>(N=88) | *p* values | APOE ε4+<br>(N=59) | APOE ε4-<br>(N=106) | *p* values |
| Cross-sectional analysis<br>(N=167) | Age, y | 53.00 (6.00) | 52.00 (11.25) | 0.36 | 52.00 (7.50) | 53.50 (9.00) | 0.16 |
|  | Sex (Female), % | 67.09% | 70.45% | 0.76 | 67.80% | 68.87% | 1.00 |
|  | Years of education | 16.00 (5.00) | 16.00 (4.25) | 0.55 | 17.00 (5.00) | 16.00 (5.00) | 0.21 |
| | APOE ε4 (carriers) % | 44.16% | 28.41% | 0.05 | - | - | - |
|  | CAIDE | 6.00 (3.00) | 5.00 (4.00) | 0.006 | 7.00 (3.00) | 5.00 (2.75) | 0.0007 |
| | | FHD+<br>(N=76) | FHD-<br>(N=68) | *p* values | APOE ε4+<br>(N=54) | APOE ε4-<br>(N=88) | *p* values |
| Longitudinal analysis<br>(N=144) | Age at baseline, y | 53.00 (6.00) | 52.00 (11.25) | 0.31 | 52.00 (7.00) | 54.00 (8.00) | 0.17 |
|  | Sex (Female), % | 67.11% | 69.12% | 0.94 | 66.67% | 68.18% | 0.99 |
|  | Years of education | 16.00 (5.00) | 17.00 (5.00) | 0.16 | 17.00 (5.00) | 16.00 (4.25) | 0.64 |
| | APOE ε4 (carriers) % | 44.59% | 30.88% | 0.13 | - | - | - |
|  | CAIDE at baseline | 6.00 (3.00) | 5.00 (4.00) | 0.004 | 7.00 (3.00) | 5.00 (3.00) | 0.0003 |

Values shown are in median (interquartile range, IQR) where applicable. *p* values were obtained from Mann-Whitney tests for continuous variables and from chi-squared tests for categorical variables. Abbreviations: FHD-, Negative family history of dementia; FHD+, positive family history of dementia; APOE ε4+, Apolipoprotein ε4 genotype positive; APOE ε4-, Apolipoprotein ε4 genotype negative. CAIDE, Cardiovascular Risk Factors, Aging and Incidence of Dementia

##### Cross-sectional effects

Baseline data were used to examine cross-sectional effects. Multiple linear regression models were used to examine the associations of global and individual network Pc with risk factors (APOE ε4 genotype, FHD and CAIDE score), each in a separate model. Age, sex, years of education, mean FD and number of brain nodes were included as covariates in all models. Network Pc showing a significant risk effect was further assessed in relation to cognitive performance using multiple linear regression models controlling for age, sex and years of education.

##### Longitudinal effects

Networks showing cross-sectional change in the presence of a risk factor were also assessed for longitudinal change over 2 years. Specifically, mean FD and number of brain nodes were first adjusted for network Pc at baseline and follow-up separately. Change scores between the two study time points were then derived to evaluate longitudinal change. Multiple linear regression models were then applied with change scores in network Pc as the dependent variable and the risk factor as the independent variable, and age at baseline, sex, and years of education as covariates. Finally, for observed significant risk-related longitudinal changes in network segregation, we performed paired t-tests to see which risk groups showed significant changes in network Pc between baseline and follow-up. We also assessed the longitudinal relationship between network segregation and cognition using multiple linear regression models. Change scores in cognition were treated as the dependent variable and change scores in network Pc as the independent variable, controlling for age at baseline, gender and years of education.

### 2.6 Validation analyses for the PREVENT-Dementia study

#### Different participant exclusion criteria based on the brain coverage in the Power atlas

To assess the impact of the chosen node retention threshold 70% for participant inclusion on the main results, we repeated the same analyses using two more stringent inclusion thresholds: 75% and 80%, i.e., excluding participants with less than 75% and 80% of the original brain nodes (node = 214) (Figure 1). The distribution of the number of remaining brain nodes in each brain network across the three thresholds is shown in Supplementary Figure 2.

#### Different parcellation schemes

To ensure that the individualised brain networks did not bias the network analyses, we used an alternative parcellation scheme (Figure 2b) that included a smaller number of key brain nodes (node = 33, spherical, 5 mm diameter) in 7 predefined brain networks (Raichle, 2011). By using this parcellation scheme, we were able to retain all brain nodes and exclude participants without full coverage of these nodes, leading to a different subset of participants compared to the main analyses (based on the 70% node retention threshold in the Power atlas) (Figure 1).

## 3 Results

### 3.1 Demographic characteristics

#### The Cam-CAN cohort

Demographic specifications of the whole cohort were summarized in Table 1.

#### The PREVENT cohort

Demographic information of the cross-sectional and longitudinal cohorts after quality control of the fMRI data based on global network coverage (Figure 2b), stratified by *APOE* ε4 genotype, and FHD, is shown in Table 2. There were no significant differences in age, sex or years of education between the groups. The frequency of the *APOE* ε4 allele was higher in the FHD+ than in the FHD-at baseline (*p* = 0.05), but did not differ significantly between the two groups for the longitudinal data. CAIDE scores were significantly higher in the FHD+ group than in the FHD-group both cross-sectionally (*p* = 0.006) and longitudinally (*p* = 0.004). Naturally, the CAIDE scores including *APOE* status were significantly higher in the *APOE* ε4+ group than in the *APOE* ε4-group, both cross-sectionally (*p* = 0.0007) and longitudinally (*p* = 0.0003) (Table 2).

3.2 Functional network segregation across the healthy adult lifespan from the Cam-CAN study

We first examined the associations (i) between global network segregation and mean cortical GMV and (ii) between global network segregation and age across the adult lifespan, to establish the efficacy of the participation coefficient (Pc) as a metric of brain health.

The multiple linear regression models showed a significant negative association between global Pc and GMV (*β* = −0.14, *p* = 0.03, Figure 3b), independent of age, sex, educational attainment, and head motion (i.e., mean FD) (Table 3). Higher global Pc (lower network segregation) was significantly associated with smaller mean cortical GMV. There was also a significant positive association between global Pc and age (*β* = 0.27, *p* < 0.0001, Figure 3c), independent of sex, educational attainment, and mean FD (Table 3). Increasing adult age was significantly associated with increased global Pc (reduced network segregation). In addition, sex was significantly associated with global Pc (*β* = −0.16, *p* = 0.02). Females had significantly lower global Pc (higher segregation) than males.

**Figure 3.**
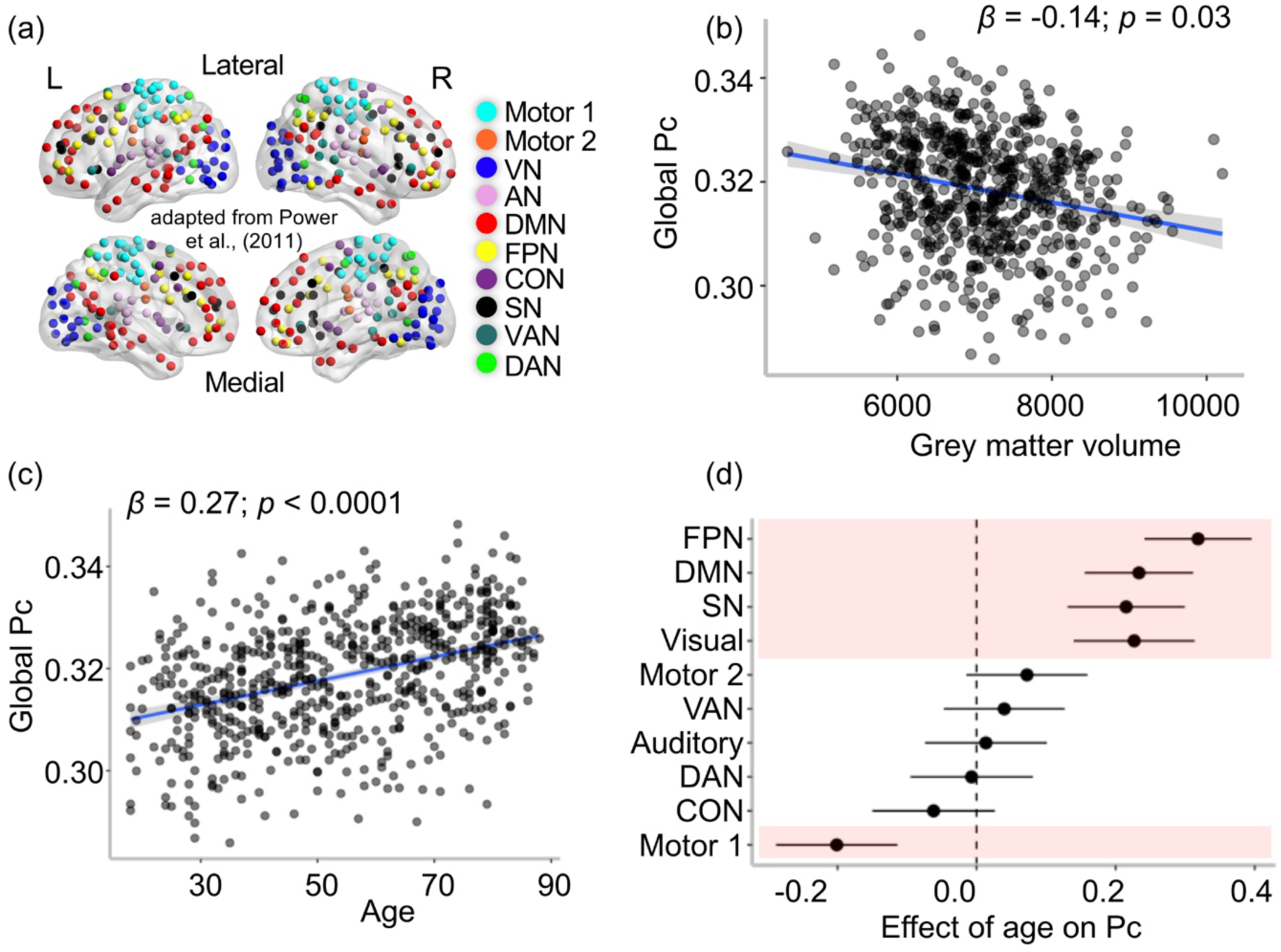
Associations between participant coefficient (Pc) with age and grey matter volume across the brain. (a) Ten comprehensive predefined networks based on Power et al. (2011). (b) Higher global Pc (loss of segregation) was significantly associated with smaller grey matter volume. (c) Higher global Pc (loss of segregation) was significantly associated with increasing age. (d) Network-specific effect of age on Pc. Standardized β coefficient (dot) and 95% confidence interval (CI, horizontal line) were shown for each network. Any CI encompasses zero (vertical dash line) represented non-significant results. Shading areas indicate significant results after Bonferroni correction across ten networks. Abbreviations: FPN, frontal-parietal control; SN, salience; DMN, default mode; VAN, ventral attention; DAN, dorsal attention; CON, cingulo-opercular control.

**Table 3.** Regression models assessing associations of global participation coefficient (Pc) with mean cortical grey matter volume (GMV) and with age, controlling for covariates.

| Dependent variables | Independent variables | $\beta$ | 95% CI | $t$ | $p$ |
| --- | --- | --- | --- | --- | --- |
| Global Pc | GMV | -0.14 | [-0.27, -0.01] | -2.17 | 0.03 |
|  | Age | 0.14 | [0.01, 0.27] | 2.12 | 0.03 |
|  | Sex | -0.13 | [-0.27, 0.01] | -1.81 | 0.07 |
|  | Education | -0.07 | [-0.14, 0.00] | -1.84 | 0.07 |
|  | Mean FD | 0.23 | [0.15, 0.31] | 5.79 | <0.0001 |
| Global Pc | Age | 0.27 | [0.19, 0.34] | 6.57 | <0.0001 |
|  | Sex | -0.16 | [-0.30, -0.03] | -2.33 | 0.02 |
|  | Education | -0.05 | [-0.12, 0.03] | -1.28 | 0.20 |
|  | Mean FD | 0.24 | [0.16, 0.32] | 6.10 | <0.0001 |

*Note: Standard coefficient $\beta$ was reported. To account for inter-individual differences in head size, grey matter volume in the first model was normalised to intracranial volume. CI = confidence interval; FD = framewise displacement.*

Network segregation is positively associated with structural brain integrity, and negatively associated with age throughout adulthood, showing that it tracks structural markers of brain health. Therefore, these findings suggest that network segregation serves as a functional marker of brain health. In the next section, we conducted network-by-network analyses of age-related changes in functional segregation.

To investigate the selective vulnerability of individual networks to healthy ageing, we assessed the associations between Pc of each of the ten brain networks (Figure 3a) and age. Pc of the DMN (*β* = 0.23, *p* < 0.0001), FPN (*β* = 0.32, *p* < 0.0001), SN (*β* = 0.22, *p* < 0.0001) and the Visual network (*β* = 0.23, *p* < 0.0001) were significantly positively associated with age (Figure 3d), independent of sex, education, and mean FD (Table 4), indicating significant age-related decreases in the functional segregation of these networks. Conversely, we observed a significant negative association between Pc of the Motor 1 network and age (*β* = - 0.20, *p* < 0.0001, Figure 3d), suggesting a significant age-related increase in functional segregation. The other networks showed no significant associations with age (Table 4). All results reported here were Bonferroni corrected.

**Table 4.**
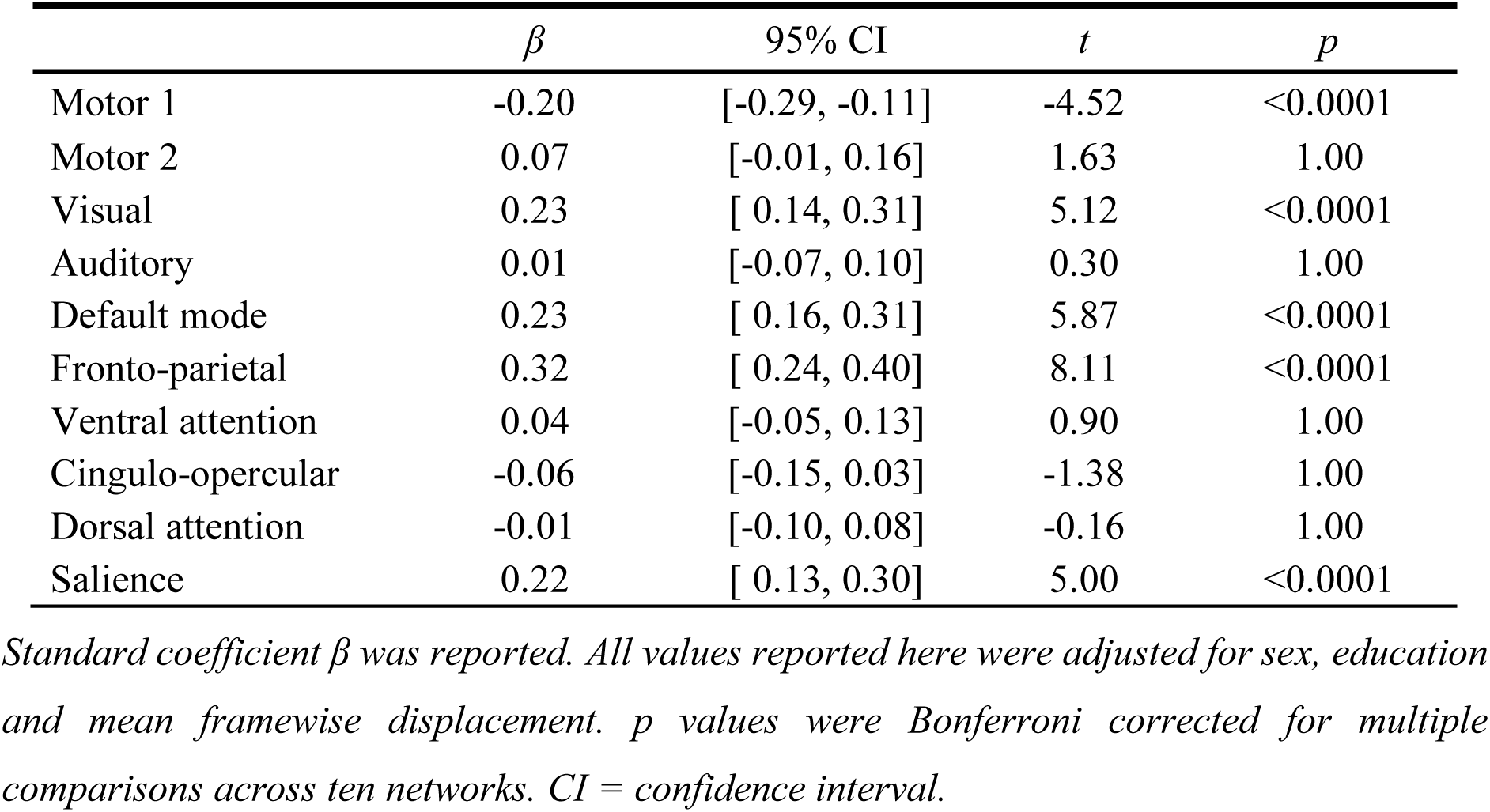
Associations of network-specific participation coefficient (Pc) with age.

The results for the three higher-order networks (FPN, DMN, SN) were consistent with our hypothesis, based on prior literature. Those networks involved in high-level cognitive processing showed the strongest age effects. However, the age effect on the functional segregation of the Motor 1 network was not predicted and adds to the existing literature. In the following sections, we investigated the effects of risk factors for late-onset AD on network segregation in midlife both cross-sectionally and longitudinally, using the PREVENT-Dementia cohort.

### 3.3 Network segregation and AD risk factors in midlife cross-sectionally

At baseline, we found a significant negative association between global Pc and *APOE* ε4 genotype (β = −0.44, *p* = 0.004), independent of age, sex, years of education, mean FD and number of brain nodes (Table 5). *APOE* ε4 carriers had significantly lower Pc (higher segregation) than ε4 non-carriers (Figure 4a). This result was further supported by validation analyses using different participant exclusion thresholds (75% and 80% retained nodes in the Power et al. (2011) atlas), and using a second brain parcellation scheme by Raichle (2011) (Supplementary Table 1). Neither family history of dementia nor the CAIDE score was significantly associated with global Pc at baseline, controlling for covariates (Supplementary Table 2).

**Figure 4.**
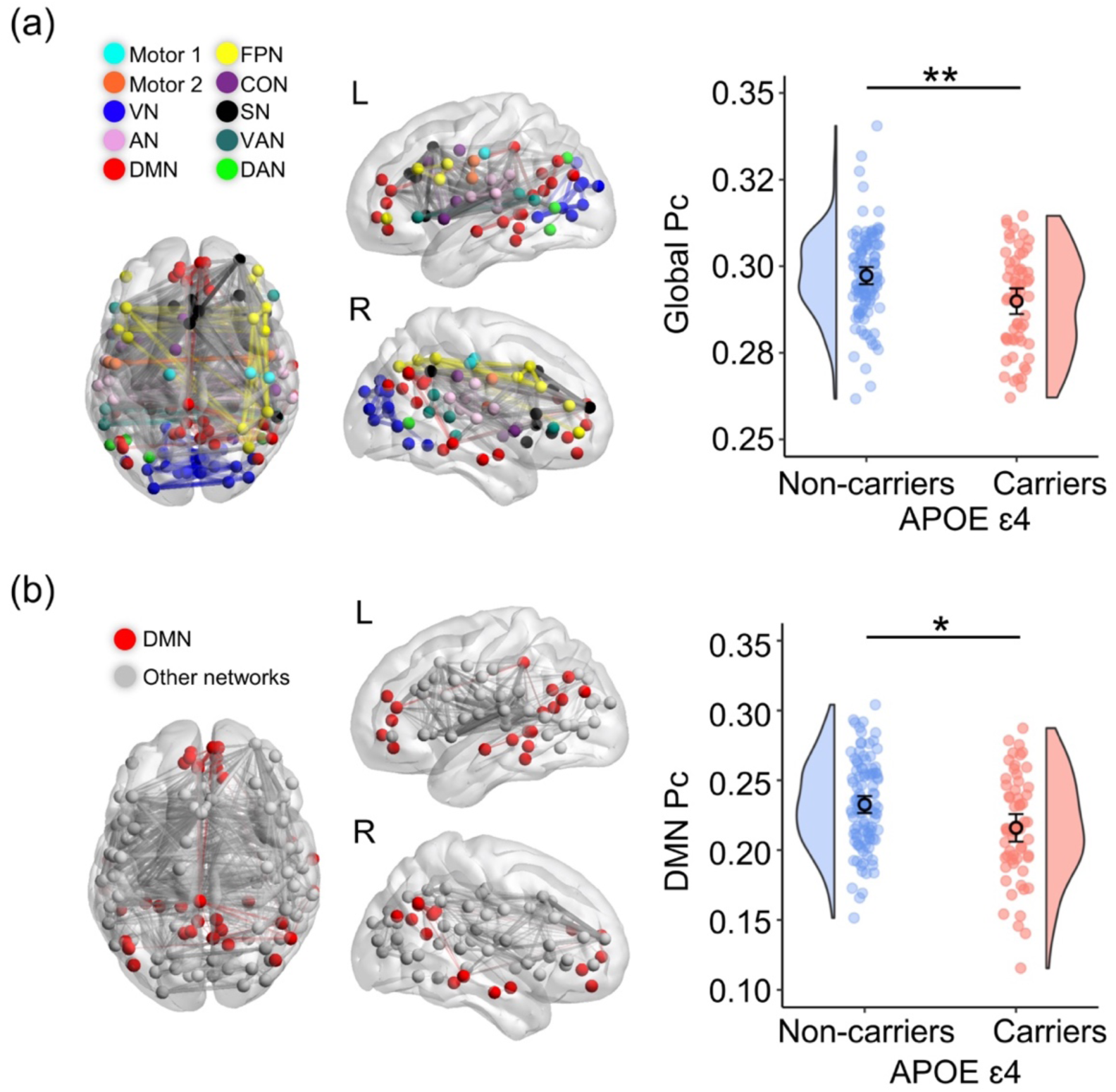
Cross-sectional associations between APOE ε4 allele and participation coefficient (Pc) of functional brain networks. Brain nodes of 10 predefined networks retained for every participant were displayed (left panel) in a 3D glass brain just for visualization purpose. Coloured lines represent the functional connectivity within particular networks (e.g., red for DMN) and grey lines represent the between network functional connectivity. (a) APOE ε4 carriers had lower global Pc (higher segregation) relative to non-carriers. (b) APOE ε4 carriers had lower Pc (higher segregation) of the default mode network (DMN) relative to non-carriers. Abbreviations: VN, visual; AN, auditory; DMN, default mode; FPN, frontal-parietal control; SN, salience; VAN, ventral attention; DAN, dorsal attention; CON, cingulo-opercular control. **p<0.005; *p<0.05, FDR corrected.

**Table 5.** Associations between the main genetic risk factor for late-onset Alzheimer’s disease apolipoprotein ε4 (APOE ε4) allele and global participation coefficient (Pc) at baseline.

| Dependent variables | Independent variables | $\beta$ | 95% CI | $t$ | $p$ |
| --- | --- | --- | --- | --- | --- |
| Global Pc | APOE $\epsilon 4$ | -0.44 | [-0.74, -0.15] | -2.94 | 0.004 |
|  | Age | -0.06 | [-0.20, 0.08] | -0.85 | 0.40 |
|  | Sex | 0.003 | [-0.30, 0.31] | 0.02 | 0.99 |
|  | Years of education | 0.05 | [-0.09, 0.20] | 0.73 | 0.47 |
|  | Mean FD | 0.25 | [0.11, 0.39] | 3.44 | 0.001 |
|  | no. of brain nodes | 0.27 | [0.13, 0.41] | 3.71 | 0.0003 |

*Standard coefficient $\beta$ was reported. CI = confidence interval. FD = framewise displacement.*

The investigation of network-specific effect of *APOE* ε4 genotype on functional segregation showed that, cross-sectionally, of the ten networks, only the DMN showed a significant negative association between *APOE* ε4 genotype and Pc at baseline (β = −0.48, *p* = 0.02, FDR correction), independent of age, sex, years of education, mean FD and number of brain nodes (Table 6). *APOE* ε4 carriers had significantly lower Pc (higher segregation) of the DMN than non-carriers (Figure 4b). This result was also supported by the validation analyses (Supplementary Table 1). Other networks did not show significant associations with *APOE* genotype (Table 6).

**Table 6.** Baseline differences in participation coefficient for 10 predefined networks (Power et al., 2011) between APOE ε4 carriers (+) and non-carriers (-).

| Network | *APOE* $\epsilon 4$ + | *APOE* $\epsilon 4$ - | $\beta$ | 91% CI | *p* |
| --- | --- | --- | --- | --- | --- |
| Motor 1 | 0.35 ± 0.01 | 0.35 ± 0.01 | 0.13 | [-0.20, 0.45] | 0.77 |
| Motor 2 | 0.33 ± 0.03 | 0.33 ± 0.03 | -0.07 | [-0.39, 0.26] | 0.87 |
| CON | 0.33 ± 0.04 | 0.34 ± 0.03 | -0.29 | [-0.61, 0.04] | 0.37 |
| AN | 0.35 ± 0.02 | 0.35 ± 0.02 | 0.08 | [-0.25, 0.40] | 0.87 |
| DMN | 0.22 ± 0.04 | 0.23 ± 0.03 | -0.48 | [-0.78, -0.18] | 0.02 |
| VN | 0.29 ± 0.03 | 0.29 ± 0.03 | 0.12 | [-0.20, 0.45] | 0.77 |
| FPN | 0.28 ± 0.03 | 0.29 ± 0.03 | -0.19 | [-0.51, 0.14] | 0.64 |
| SN | 0.31 ± 0.04 | 0.32 ± 0.03 | -0.26 | [-0.58, 0.06] | 0.37 |
| VAN | 0.32 ± 0.03 | 0.32 ± 0.03 | -0.04 | [-0.36, 0.29] | 0.90 |
| DAN | 0.33 ± 0.02 | 0.33 ± 0.02 | -0.02 | [-0.34, 0.30] | 0.90 |

*The shown values are mean ± standard deviation with the standardized coefficient ( $\beta$ ) and 95% confidence interval (CI) from the linear regression models while controlling for age, sex, years of education, mean FD and the number of brain nodes. Abbreviations: AN, auditory; VN, visual; DMN, default mode; FPN, frontal-parietal control; VAN, ventral attention; CON, cingulo-opercular control; DAN, dorsal attention; SN, salience network. *p* values were false discovery rate (FDR) corrected.*

### 3.4 Longitudinal effects of AD risk factors on network segregation in midlife

Then, we assessed the associations of longitudinal changes in global and DMN segregation with *APOE* ε4 allele carriership. We found a trend positive association between *APOE* ε4 allele and longitudinal change in global Pc (! = 0.32, *p* = 0.06, Table 7), independent of baseline age, sex and years of education. Paired *t* tests showed no significant longitudinal change in global Pc for either *APOE* ε4 carriers or non-carriers (Figure 5a). There was a significant positive association between baseline age and longitudinal change in global Pc (! = 0.26, *p* = 0.002, Table 7). Older age at baseline was significantly associated with greater longitudinal increase in global Pc (loss of segregation) (Figure 6a).

**Figure 5.**
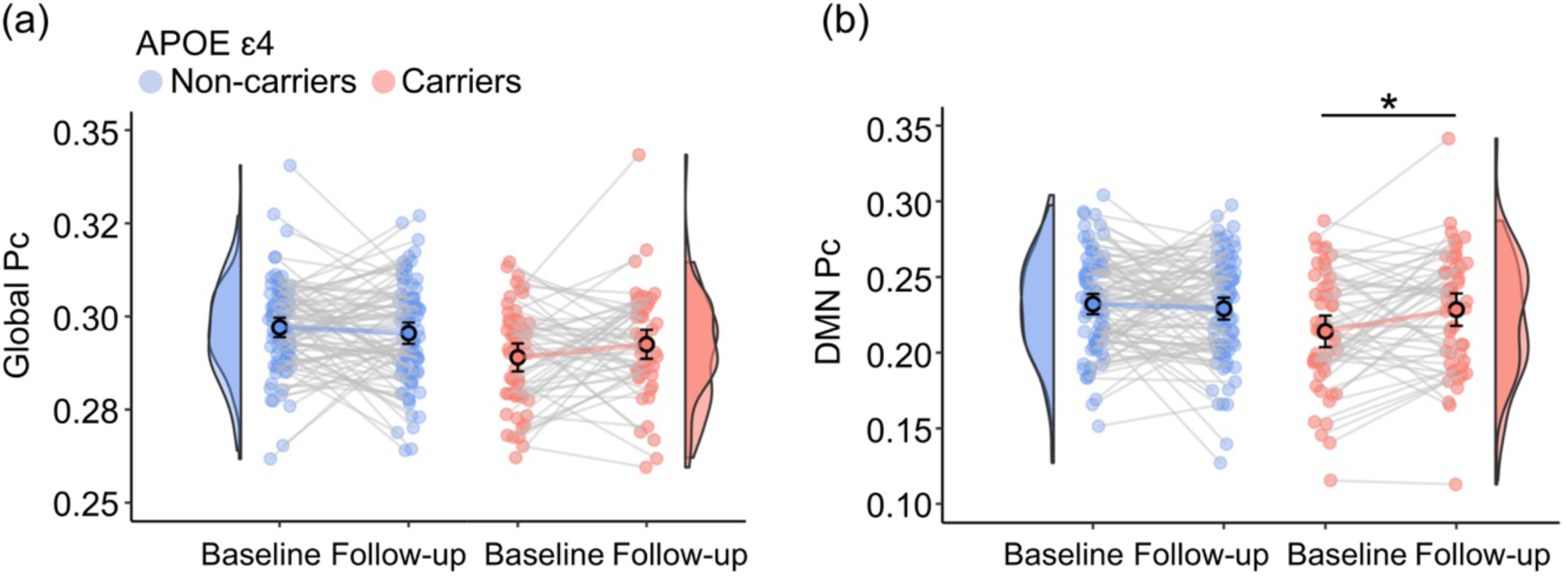
Longitudinal associations between APOE ε4 allele and participation coefficient (Pc) of functional brain networks over 2 years. (a) No significant change in global Pc for either groups. (b) APOE ε4 carriers only showed significantly increased Pc of the default mode network (DMN). *p<0.05, FDR corrected.

**Figure 6.**
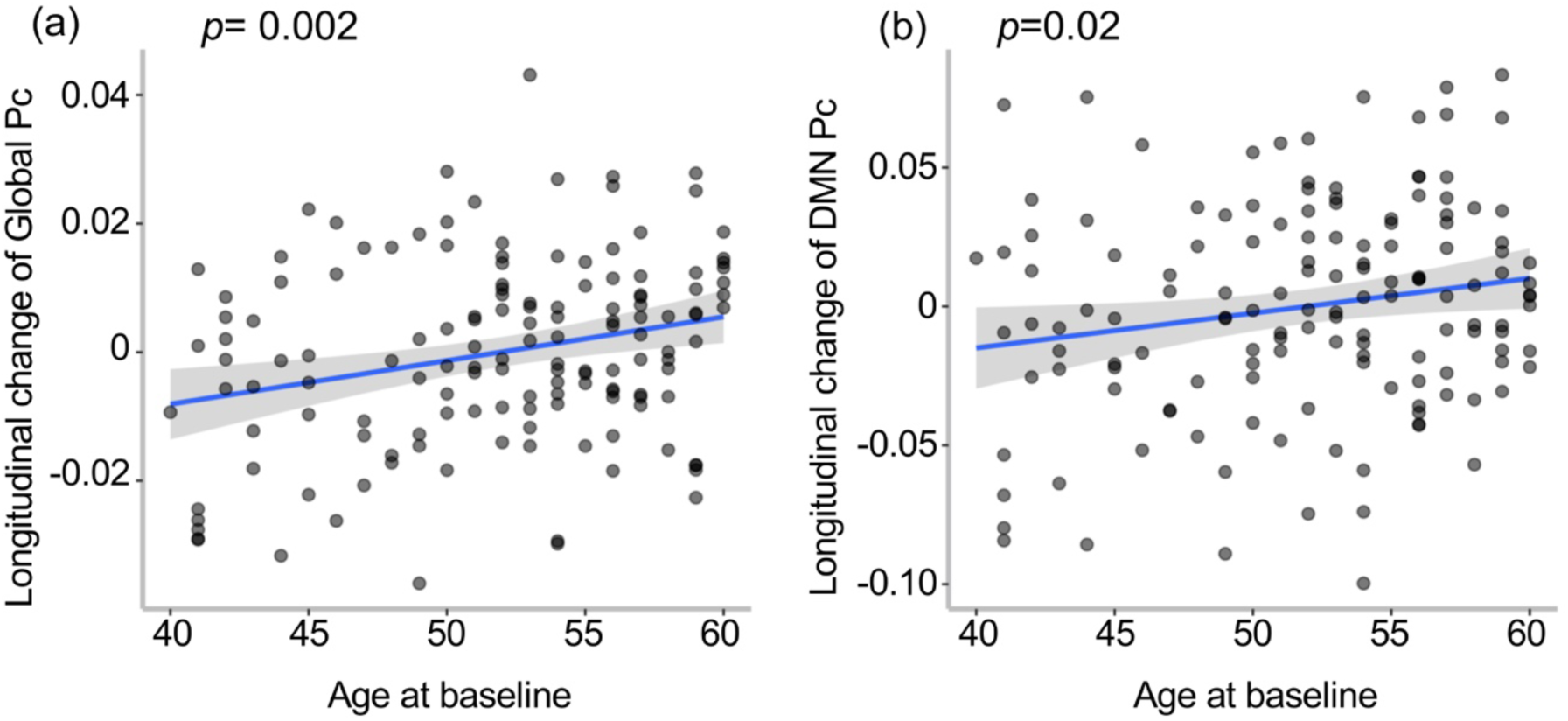
Associations between baseline age and longitudinal change of the participation coefficient (Pc). (a) increased age at baseline was associated with longitudinal increase of the global Pc (loss of segregation). (b) increased age at baseline was associated with longitudinal increase of the default mode network (DMN) Pc (loss of segregation).

**Table 7.** Longitudinal changes (over 2 years) in global and default model network (DMN) participation coefficient in relation to apolipoprotein ε4 (APOE ε4) allele.

| Dependent variables | Independent variables | $\beta$ | 95% CI | $t$ | $p$ |
| --- | --- | --- | --- | --- | --- |
| $\Delta$ Global Pc | $APOE \epsilon 4$ | 0.32 | [-0.01, 0.65] | 1.90 | 0.06 |
|  | Age at baseline | 0.26 | [0.10, 0.43] | 3.20 | 0.002 |
|  | Sex | 0.13 | [-0.21, 0.48] | 0.76 | 0.45 |
|  | Years of education | -0.09 | [-0.25, 0.07] | -1.08 | 0.28 |
| $\Delta$ DMN Pc | $APOE \epsilon 4$ | 0.46 | [0.12, 0.79] | 2.70 | 0.008 |
|  | Age at baseline | 0.19 | [0.03, 0.36] | 2.34 | 0.02 |
|  | Sex | 0.20 | [-0.15, 0.54] | 1.12 | 0.26 |
|  | Years of education | -0.03 | [-0.19, 0.13] | -0.37 | 0.71 |

Standard coefficient $\beta$ was reported. CI = confidence interval. $\Delta$ = follow-up – baseline.

Critically, we found a significant positive association between *APOE* ε4 genotype and longitudinal change in DMN Pc (! = 0.46, *p* = 0.008, Table 7), independent of baseline age, sex and years of education. Paired *t* tests showed that only *APOE* ε4 carriers had a significant increase in DMN Pc (loss of network segregation) over 2 years (*t* = −2.80, *p* = 0.007, Figure 5b), and *APOE* ε4 non-carriers showed no significant longitudinal change. We also observed a significant positive association between the DMN Pc and baseline age (! = 0.19, *p* = 0.02, Table 7). Older age at baseline was significantly associated with greater longitudinal increase in DMN Pc (loss of segregation) (Figure 6b).

3.5 Network segregation and cognitive performance, cross-sectionally and longitudinally

Finally, we investigated the association between global and DMN segregation with cognitive performance. Cross-sectionally, there was a significant negative association between global Pc and episodic and relational memory at baseline, independent of age, sex, and years of education (*β* = −0.19, *p* = 0.009). Higher global Pc (lower segregation) was significantly associated with worse cognition (Figure 7a). The other two cognitive domains did not show significant relationships with global Pc (Table 8). Higher Pc of the DMN (lower segregation) was also significantly associated with worse episodic and relational memory at baseline (! = −0.17, *p* = 0.02, Figure 7b), but not the other two domains (Table 9), independent of age, sex, and years of education. Longitudinally, there were no significant associations between the change scores in any of the three cognitive domains over the 2 years and the change scores in either the global Pc or the DMN Pc.

**Figure 7.**
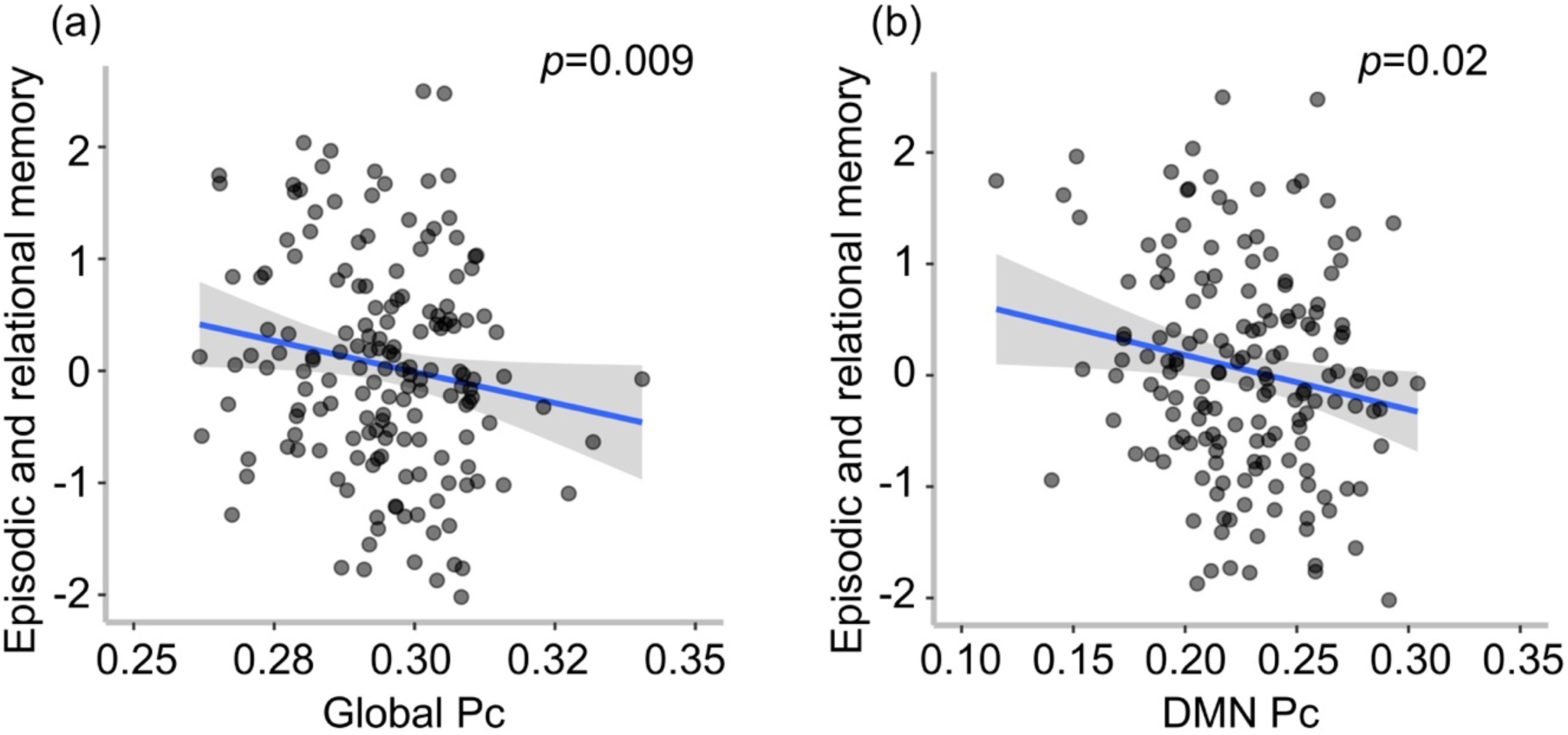
Cross-sectional relationships between participation coefficient (Pc) and cognitive performance from the PREVENT-Dementia study. (a) Higher global Pc (lower network segregation) and (b) higher DMN Pc (lower network segregation) were significantly associated with worse episodic and relational memory at baseline.

**Table 8.** Associations between global participation coefficient (Pc) and cognition in cognitively unimpaired middle-aged adults from the PREVENT-dementia study at baseline.

| Dependent variables | Independent variables | $\beta$ | 95% CI | $t$ | $p$ |
| --- | --- | --- | --- | --- | --- |
| Episodic and relational episodic memory | Global Pc | -0.19 | [-0.33, -0.05] | -2.64 | 0.009 |
|  | Age | -0.08 | [-0.22, 0.06] | -1.12 | 0.27 |
|  | Sex | 0.23 | [-0.08, 0.54] | 1.48 | 0.14 |
|  | Years of education | 0.35 | [0.20, 0.49] | 4.81 | <0.0001 |
| Working and short-term (single-feature) memory | Global Pc | 0.08 | [-0.07, 0.23] | 1.02 | 0.31 |
|  | Age | -0.06 | [-0.21, 0.10] | -0.73 | 0.46 |
|  | Sex | -0.25 | [-0.58, 0.08] | -1.47 | 0.14 |
|  | Years of education | 0.15 | [0.00, 0.30] | 1.91 | 0.06 |
| Verbal and visuospatial functions, and short-term (conjunctive) memory | Global Pc | -0.06 | [-0.21, 0.09] | -0.81 | 0.42 |
|  | Age | -0.20 | [-0.35, -0.05] | -2.56 | 0.01 |
|  | Sex | 0.18 | [-0.14, 0.51] | 1.11 | 0.27 |
|  | Years of education | -0.01 | [-0.16, 0.14] | -0.14 | 0.89 |

*Note: Standard coefficient $\beta$ was reported. CI = confidence interval.*

**Table 9.** Associations of the default model network (DMN) participation coefficient (Pc) and cognition in cognitively unimpaired middle-aged adults from the PREVENT-dementia study at baseline.

| Dependent variables | Independent variables | $\beta$ | 95% CI | $t$ | $p$ |
| --- | --- | --- | --- | --- | --- |
| Episodic and relational episodic memory | DMN Pc | -0.17 | [-0.32, -0.03] | -2.33 | 0.02 |
|  | Age | -0.10 | [-0.24, 0.05] | -1.35 | 0.18 |
|  | Sex | 0.17 | [-0.14, 0.48] | 1.07 | 0.29 |
|  | Years of education | 0.33 | [0.19, 0.47] | 4.56 | <0.0001 |
| Working and short-term (single-feature) memory | DMN Pc | 0.08 | [-0.08, 0.24] | 1.00 | 0.32 |
|  | Age | -0.05 | [-0.20, 0.11] | -0.61 | 0.54 |
|  | Sex | -0.22 | [-0.55, 0.12] | -1.28 | 0.20 |
|  | Years of education | 0.16 | [ 0.00, 0.31] | 2.00 | 0.05 |
| Verbal and visuospatial functions, and short-term (conjunctive) memory | DMN Pc | -0.12 | [-0.28, 0.03] | -1.55 | 0.12 |
|  | Age | -0.21 | [-0.37, -0.06] | -2.76 | 0.007 |
|  | Sex | 0.14 | [-0.19, 0.47] | 0.83 | 0.41 |
|  | Years of education | -0.02 | [-0.17, 0.13] | -0.26 | 0.80 |

*Note: Standard coefficient $\beta$ was reported. CI = confidence interval.*

## 4 Discussion

Functional segregation, a summary measure of brain network organisation, is an emerging measure of brain health in both normal and pathological ageing processes (Brier et al., 2014; Wig, 2017). In this study, we investigated the vulnerability of brain networks to loss of segregation during healthy adult lifespan and in cognitively healthy midlife individuals at risk for late-onset AD, as well as the association between segregation loss and cognition in midlife. First, at the global brain level, we showed that: (i) greater network segregation was significantly associated with greater mean cortical GMV, a putative measure of brain health, consistent with previous work (Kong et al., 2020), and (ii) network segregation was significantly reduced with increasing adult age, suggesting that brain networks become less segregated or more diffuse during healthy ageing, consistent with previous studies (Chan et al., 2014; Wig, 2017). The second relationship was further supported by a novel result from the midlife cohort of the PREVENT-Dementia study. Older age at baseline was significantly associated with reduced network segregation over two years in midlife. Taken together, these findings support that functional network segregation, in particular the Pc metric, may serve as a proxy for brain health, and could be useful for detecting early changes in individuals at risk for Alzheimer’s disease.

The second novel finding of this study was that females had higher network segregation than males across the adult lifespan, independent of age and educational attainment. There is currently a lack of knowledge about sex differences in network segregation. While one recent study examined the effect of sex on network segregation throughout adulthood, the results show no sex differences (Ballard et al., 2022). A possible explanation for this discrepancy could be a different metric of functional segregation used in that study compared to the current one. This new finding suggests that it is important to consider sex differences when investigating network segregation. Therefore, the results of the present study regarding age- or AD risk-related changes were all controlled for the sex effect. Another potential implication for future research is to determine the role of sex in age- or other AD risk-related changes in network segregation (Ballard et al., 2022), which may help to explain sex differences in the prevalence and progression of Alzheimer’s disease.

The investigation of network-specific age-related changes in functional segregation across the adult lifespan revealed significant changes in 5/10 networks, including three high-order networks (DMN, FPN and SN) and two sensorimotor networks (Visual and Motor 1). Previous studies addressing this research question have yielded inconsistent findings regarding which networks are susceptible to segregation loss across the adult lifespan (Ballard et al., 2022; Cassady et al., 2019; Chan et al., 2014), possibly due to differences in statistical power associated with different sample sizes, correction methods for multiple comparisons to control for false positive rates, and measures of network segregation (Varangis et al., 2019). Despite these differences in study design, the present study, together with previous studies (Ballard et al., 2022; Chan et al., 2014; Chong et al., 2019; Grady et al., 2016; Malagurski et al., 2020; Ng et al., 2018), provides further evidence for age-related declines in functional segregation of high-order networks, supporting their vulnerability during healthy ageing.

Contradicting previous studies (Ballard et al., 2022; Cassady et al., 2020; Cassady et al., 2019; Chan et al., 2014; Manza et al., 2020), the present study found an age-related increase in functional segregation of the Motor 1 network. Although this change in direction is counterintuitive, it is consistent with some previous studies showing increased functional connectivity within the somatomotor network throughout adulthood (He et al., 2017; Mathys et al., 2014; Song et al., 2014; Tomasi & Volkow, 2012), suggesting a compensatory role of such increases in response to declining motor function. A possible reason for the discrepancy between previous studies and the current study is that there may be a non-linear relationship between age and functional segregation of the motor network across the adult lifespan (Varangis et al., 2019), leading to mixed findings when examining their linear relationship (Jockwitz & Caspers, 2021). Further studies are needed to test this hypothesis. Nevertheless, the networks that showed significant changes in functional segregation with age, particularly the high-order networks, have important implications for predicting changes in early AD processes in midlife.

Critically, the third novel finding of this study was that cross-sectionally, cognitively healthy middle-aged adults carrying an *APOE* ε4 allele showed greater global network segregation compared to non-carriers. This change in direction is consistent with previous studies of the same and other similar midlife cohorts showing better cognition (Deng et al., 2022; Gharbi-Meliani et al., 2021; Ritchie et al., 2017; Zokaei et al., 2020), cerebral hyperperfusion (Dounavi et al., 2021; Mak et al., 2021; McKiernan et al., 2020) and hyperconnectivity within the DMN (Cacciaglia et al., 2022; Westlye et al., 2011) in *APOE* ε4 carriers than in non-carriers. Importantly, greater network segregation, particularly in the DMN, was significantly associated with better episodic and relational memory cross-sectionally in this midlife cohort. The DMN has been widely recognized for its critical role in episodic memory (Dickerson & Sperling, 2009), and our novel finding further suggests that maintaining functional segregation of the DMN is crucial for better episodic and relational memory in midlife. However, the longitudinal analysis did not show a significant association between change in network segregation and change in cognition over two years, which may be due to the relatively young age range of the sample, leading to small variations in brain function and cognition changes over such a short follow-up period at the whole group level.

Furthermore, of the ten networks, only the DMN showed an effect of *APOE* genotype, with ε4 carriers showing higher DMN segregation than non-carriers at baseline. The DMN has been shown to be vulnerable to AD pathology, and its functional connectivity has been extensively investigated at different stages of AD (Greicius et al., 2004; Habib et al., 2017; Koch et al., 2012; Kucikova et al., 2021; Sorg et al., 2007). In preclinical AD, beta-amyloid (Aβ) deposition strongly overlaps with DMN regions (Buckner et al., 2005; Palmqvist et al., 2017) and is inversely associated with DMN functional connectivity in cognitively healthy older adults (Mormino et al., 2011; Palmqvist et al., 2017; Sheline et al., 2010). DMN functional connectivity has also been found to be disrupted in patients with MCI (Gili et al., 2011) and AD (Gour et al., 2014; Grieder et al., 2018), and to track disease progression and conversion from MCI to AD (Brier et al., 2012; Damoiseaux et al., 2012; Petrella et al., 2011). These findings highlight the vulnerability of the DMN functional connectivity across the AD spectrum. Our novel finding further suggests that the DMN is vulnerable to changes in functional organization in middle-aged individuals who are currently cognitively healthy but at genetic risk for late-onset AD.

Moreover, *APOE* ε4 carriers, but not non-carriers, showed a significant loss of network segregation in the DMN over two years. To the best of our knowledge, this is the first study to show a prominent decline in functional segregation of the DMN with ageing in cognitively healthy individuals at risk for late-onset AD in midlife. A greater loss of network segregation with ageing, particularly in high-order networks (i.e., DMN, FPN and SN), was previously shown in cognitively unimpaired older *APOE* ε4 carriers compared to non-carriers (Ng et al., 2018). Furthermore, accelerated age-related decline in network segregation was predictive of dementia severity (Chan et al., 2021). Our novel finding extends these previous findings by demonstrating an accelerated decline in functional segregation in individuals at higher risk of late-onset AD in midlife, decades before the onset of clinical manifestations of dementia.

Taken together, the present study demonstrates the impact of the major genetic risk for sporadic AD in the Caucasian population on functional brain network organization in midlife, and further uncovers a previously unknown trajectory: stronger functional segregation of *APOE* ε4 carriers cross-sectionally, followed by a pronounced age-related loss of segregation longitudinally, relative to non-carriers. These results lend some support to a recent proposal on the impact of *APOE* ε4 on AD biomarker progression trajectories (Koelewijn et al., 2019), which hypothesizes a dichotomised effect of *APOE* ε4 on functional brain biomarkers, i.e., hyper-expression, e.g., hyperactivity / hyperconnectivity, in late young adulthood (i.e., from the 30s onwards), and hypo-expression, e.g., hypoconnectivity in later life. In support of this hypothesis, Koelewijn et al. (2019) found significantly higher functional connectivity of brain networks as measured with MEG, particularly in the DMN, in young *APOE* ɛ4 carriers (age: 24.5 ± 5.4 years) compared to age-matched non-carriers (Koelewijn et al., 2019), but significantly reduced functional connectivity in clinical AD patients (age: 67-89 years) compared to age-matched healthy controls (Koelewijn et al., 2017). However, their comparisons were cross-sectional, making it impossible to draw conclusions about the longitudinal course of changes. In addition, the participants included in their study did not cover the midlife stage (Koelewijn et al., 2017; Koelewijn et al., 2019). Our findings on longitudinal changes, with a specific focus on midlife, therefore significantly advance this field. Why does the *APOE* genotype show a dichotomised expression in brain function across the lifespan? One possible explanation is the antagonistic pleiotropy hypothesis of aging (Williams, 1957), which proposes that deleterious genes, such as the *APOE* ε4 gene allele, have survived through evolution because they may confer an advantage, early in life when humans are reproductively fit.

In conclusion, this study provides evidence for selective vulnerabilities of brain networks to disruption in network organization during healthy ageing and in cognitively healthy midlife individuals at risk for late-onset AD. In particular, three high-order networks (i.e., the DMN, FPN, and SN) and two sensorimotor networks (i.e., the Visual and Motor 1 networks) were vulnerable to the age effect across the healthy adult lifespan. Of these networks, the DMN was particularly vulnerable to AD risk in midlife. *APOE* ε4 genotype was significantly associated with altered functional brain network organization in cognitively healthy individuals in midlife, an estimated 23 years from symptoms onset. Higher network segregation cross-sectionally was accompanied by loss of segregation longitudinally only in *APOE* ε4 carriers. These novel findings suggest that functional network segregation may constitute a novel and early substrate for the impact of genetic AD risk on the brain in midlife and thus have implications for the early detection and intervention in AD.

## Methodological considerations

The resting state fMRI data from the PREVENT-Dementia research programme presents inadequate brain coverage and inconsistent scanning angles that prevent whole brain parcellation for network analyses. To address this limitation, we adapted a comprehensive brain map (Power et al., 2011) based on individual-specific brain coverage and constructed an individualised brain map for each participant, that maximises the number of participants for the network analyses. In addition, we also replicated the findings using a different brain atlas (Raichle, 2011) that contains a smaller number of brain nodes. This ensures the same constructed brain network for each participant. Finally, we also tested the main findings using two different thresholds for participant inclusion (for details please see SI). The results were robust to these different analyses.

## Data Availability

All data produced in the present study are available upon reasonable request to the authors

## Acknowledgement

F.D. was funded by the Provost PhD Award Scheme from Trinity College Dublin, to L.N. L.N. was also funded by a L’Oréal-UNESCO for Women in Science International Rising Talent Award, the Welcome Trust Institutional Strategic Support grant, and the Global Brain Health Institute Project Grant.

The PREVENT-Dementia study is supported by the UK Alzheimer’s Society (grant numbers 178 and 264), the US Alzheimer’s Association (grant number TriBEKa-17-519007) and philanthropic donations.

The Prevent Dementia study at the Dublin site is also part supported by a project grant from GBHI and philanthropic donations from Trinity Foundation.

We thank all PREVENT-Dementia participants for their enthusiastic participation in this study. We also thank the Research Delivery service at West London NHS Trust and the Wolfson Clinical Imaging Facility at Imperial College London for their support in running the study.

