## Supporting Information File for "Genetic risk of late-onset Alzheimer’s disease is associated with longitudinal loss of functional brain network segregation in middle-aged cognitively healthy individuals: The PREVENT-Dementia Study"

**Title**

**The PREVENT-Dementia study**

**Cognitive variables:**

The eleven cognitive summary variables from the COGNITO battery were:

1. Working memory: the simultaneous presentation of auditory and visual attention tasks assessed by subtracting the time taken in milliseconds on this double task from the visual task alone.
2. Working memory: the simultaneous presentation of auditory and visual attention tasks assessed by total number of correct answers for visual form recognition on this double task.
3. Narrative recall: total number of correct elements on immediate recall of a story with a temporal progression requiring attention to macrostructure.
4. Description recall: total number of correct elements recalled of a description without thematic progression requiring attention to microstructure and recall of spatial location. The narrative and description recall are similar in terms of word frequency in the language and syntactic structure.
5. Implicit memory: difference in the number of steps in the progressive build-up of names on the screen required for recognition between names never seen and number of names previously learnt in an immediate recall task.
6. Name-face association: number of faces recognized after a delay from a series of 18 faces of which 9 have been previously shown with their corresponding names.
7. Form perception: number of correct answers in the matching of complex forms to a multiple-choice array.
8. Form perception speed: mean time taken in milliseconds for each trial.
9. Phoneme comprehension: number of correct responses in the matching of a word with an image presented as part of a multiple-choice array including semantic, morphological and phonetic distractors.
10. Phoneme comprehension speed: mean time taken in milliseconds to perform.
11. Verbal fluency: total sum of the number of words generated in 30s using both a semantic (vegetables) and phonemic (letter P) cue.

The two summary variables from the Visual Short-term Memory Binding task were:

12. Visual short-term memory binding test (shape only condition): percentage of correct recognition of the shape of presented visual stimuli after a short period of retention.
13. Visual short-term memory binding test (shape-color binding condition): percentage of correct recognition of combinations of shape and color of presented visual stimuli after a short period of retention.

**Functional brain network construction**

*Individualised brain network*

To overcome the inadequate field of view (FOV) and inconsistent scanning angles of the fMRI data from the PREVENT-Dementia research programme, we adapted a comprehensive brain map (Power et al., 2011) to the specific FOVs of the participants. The procedure was as follows: 1) create a brain mask for each participant; 2) multiply the individual brain mask by the original brain map (Power et al., 2011) to obtain the individualised brain map (Figure 2b); 3) set a threshold to exclude brain nodes of small size on the individualised brain map. The initial number of voxels for each brain node on the original brain map is 81. We excluded brain nodes on the individualised brain map where the number of remaining voxels was less than 80% of the initial number (80%*81=64).

*An alternative parcellation scheme*

To ensure that the network analyses were not biased by the individualised brain maps, we used an alternative parcellation scheme with fewer (node = 33 versus node = 214 in Power atlas) but key brain regions (Raichle, 2011). This parcellation method is theoretically based on a meta-analysis of seven brain networks identified in resting-state studies, and has also been used to investigate functional connectivity in different populations in our previous work (Deng et al., 2023; Hu et al., 2022; Naci et al., 2018).

**Framewise displacement calculation**

Differentiating head realignment parameters across frames yields a six dimensional timeseries that represents instantaneous head motion, which can then be summarised as a scalar quantity, framewise displacement (FD), using the formula (Equation (1)). Specifically, this measure was calculated as the sum of the absolute values of the derivatives of the 6 realignment parameters (Power et al., 2012). Rotational displacements were converted from degrees to millimetres by calculating the displacement on the surface of a sphere with a radius of 50 mm, which is approximately the mean distance from the cerebral cortex to the centre of the head.

$$\boldsymbol{FD}_{\boldsymbol{i}}\boldsymbol{=}\left| \boldsymbol{\Delta}\boldsymbol{d}_{\boldsymbol{ix}} \right|\boldsymbol{+}\left| \boldsymbol{\Delta}\boldsymbol{d}_{\boldsymbol{iy}} \right|\boldsymbol{+}\left| \boldsymbol{\Delta}\boldsymbol{d}_{\boldsymbol{iz}} \right|\boldsymbol{+}\left| \boldsymbol{\Delta}\boldsymbol{\alpha}_{\boldsymbol{i}} \right|\boldsymbol{+}\left| \boldsymbol{\Delta}\boldsymbol{\beta}_{\boldsymbol{i}} \right|\boldsymbol{+}\left| \boldsymbol{\Delta}\boldsymbol{\gamma}_{\boldsymbol{i}} \right|\boldsymbol{(1)}$$

Where $\Delta\boldsymbol{d}_{\boldsymbol{ix}}\boldsymbol{=}\boldsymbol{d}_{\left( \boldsymbol{i-1} \right)\boldsymbol{x}}\boldsymbol{-}\boldsymbol{d}_{\boldsymbol{ix}}$ and similarly for the other rigid body parameters $\left[ \boldsymbol{d}_{\boldsymbol{iy}}\boldsymbol{,}\boldsymbol{d}_{\boldsymbol{iz}}\boldsymbol{,}\boldsymbol{\alpha}_{\boldsymbol{i}}\boldsymbol{,}\boldsymbol{\beta}_{\boldsymbol{i}}\boldsymbol{,}\boldsymbol{\gamma}_{\boldsymbol{i}} \right]$**.**

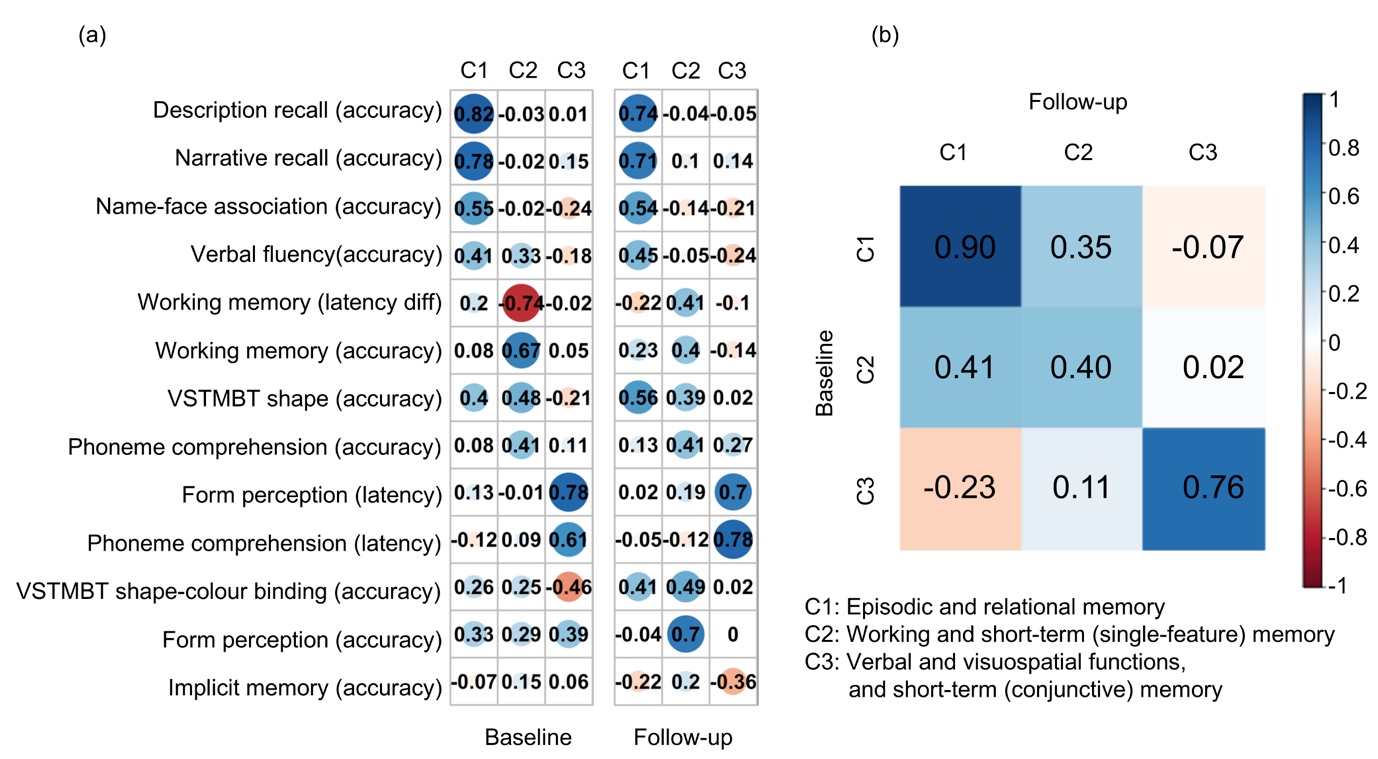

Supplementary Figure 1. Component (C) coefficients and Tucker's congruence coefficients between baseline and follow-up. (a) The coefficients for 13 original cognitive measures (in rows) on the three components (in columns). A larger absolute coefficient (darker colors and larger solid circles) represents a closer relationship between the cognitive measure and corresponding component. Cool/warm colors represent the positive/negative relationships between cognitive measures and components. (b) Similarities among the components across baseline (in rows) and follow-up (in columns) were measured by Tucker's congruence coefficients. The diagonal values represented the similarities for each of the components with itself across time and off diagonal values indicated the similarities for each of the components with the other two components across time. Cool/warm colors represent the positive/negative relationships with darker colors representing correlation magnitude, as shown in the color-bar scale. Abbreviations: VSTMBT, visual short-term memory binding test; diff, difference.

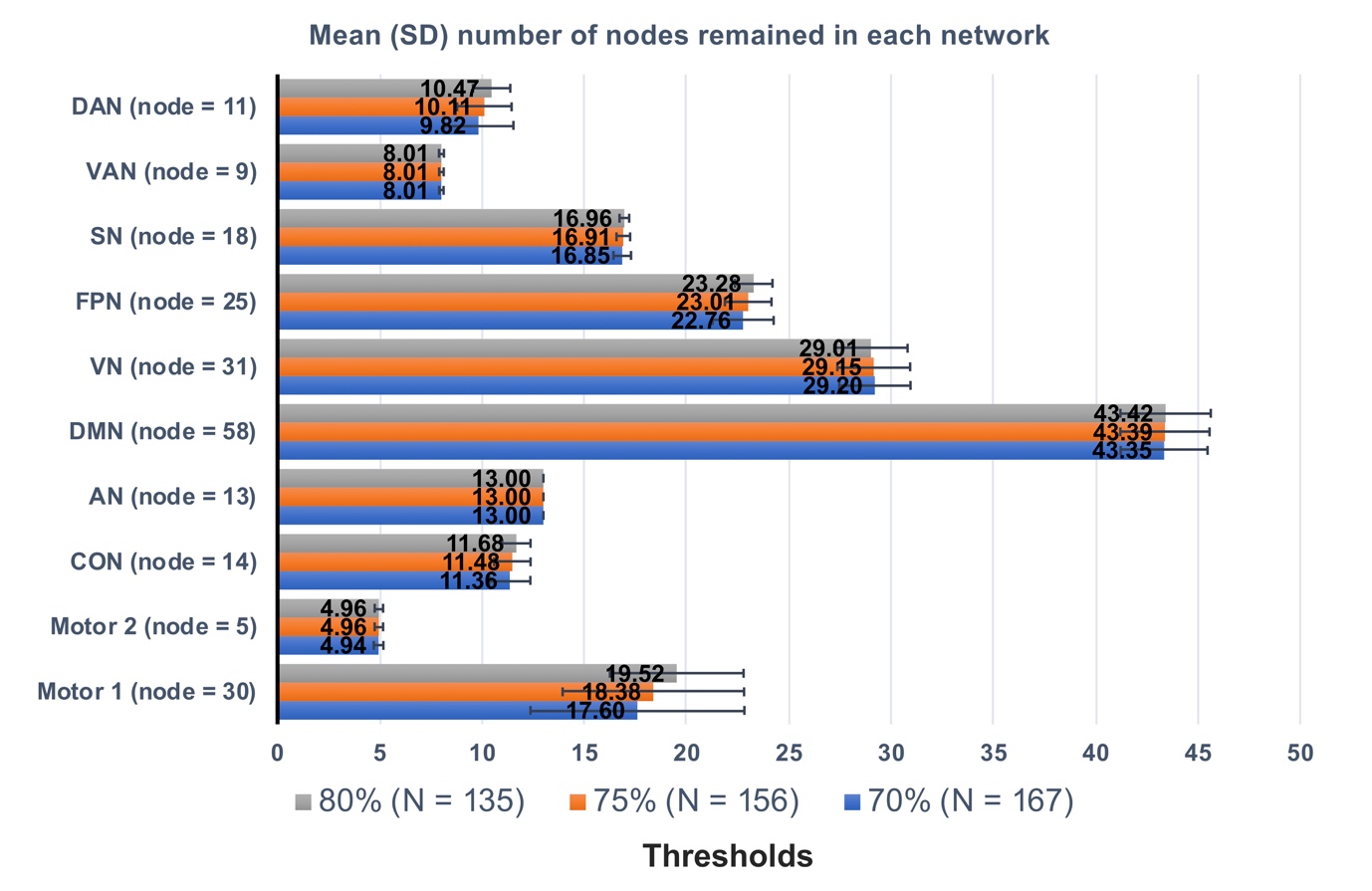

Supplementary Figure 2. The distribution of the remained brain nodes in each network based on the Power atlas (Power et al., 2011) across the three thresholds, i.e., 70%, 75% and 80%, that we used to exclude/include participants (N = the number of included participants under each threshold). The number of remained brain nodes were presented in the x axis, and the 10 different networks were shown in the y axis (node = the initial number of nodes in each network). The bar (error bar) indicates the mean (standard deviation, SD) number of nodes remained in the specific cohort after the threshold exclusion criteria. Abbreviations: VN, visual; AN, auditory; DMN, default mode; FPN, frontal-parietal control; SN, salience; VAN, ventral attention; DAN, dorsal attention; CON, cingulo-opercular control.

Supplementary Table 1 Baseline differences in the global and the default model network (DMN) participation coefficient (Pc) between *APOE* ε4 carriers (+) and non-carriers (-) for different parcellation scheme and different participant exclusion thresholds (75% and 80%) based on the number of remained brain nodes in the Power atlas.

|  | | | Global Pc | DMN Pc |
| --- | --- | --- | --- | --- |
| Power Atlas | 70% (N=167) | *APOE* ε4 + (N=59) | 0.29 ± 0.01 | 0.22 ± 0.04 |
|  |  | *APOE* ε4 - (N=106) | 0.30 ± 0.01 | 0.23 ± 0.03 |
|  |  | *p* value | 0.004 | 0.02 |
|  | 75% (N=156) | *APOE* ε4 + (N=53) | 0.29 ± 0.01 | 0.22 ± 0.04 |
|  |  | *APOE* ε4 - (N=101) | 0.30 ± 0.01 | 0.23 ± 0.03 |
|  |  | *p* value | 0.002 | 0.001 |
|  | 80% (N=135) | *APOE* ε4 + (N=45) | 0.29 ± 0.01 | 0.21 ± 0.04 |
|  |  | *APOE* ε4 - (N=88) | 0.30 ± 0.01 | 0.23 ± 0.03 |
|  |  | *p* value | 0.0004 | 0.0007 |
|  | The same participants as in Raichle Atlas (N=136) | *APOE* ε4 + (N=46) | 0.29 ± 0.01 | 0.22 ± 0.04 |
|  |  | *APOE* ε4 - (N=88) | 0.30 ± 0.01 | 0.23 ± 0.03 |
|  |  | *p* value | 0.03 | 0.02 |
| Raichle Atlas (N=136) | | *APOE* ε4 + (N=46) | 0.24 ± 0.02 | 0.17 ± 0.05 |
|  |  | *APOE* ε4 - (N=88) | 0.24 ± 0.01 | 0.18 ± 0.05 |
|  |  | *p* value | 0.009 | 0.09 |

The shown values are mean ± standard deviation; N=number of participants included in the particular criteria; 2 participants without APOE ε4 information.

Supplementary Table 2 Associations between the risk factors for late-onset Alzheimer’s disease [family history of dementia (FHD), and Cardiovascular Risk Factors, Aging, and Incidence of Dementia (CAIDE) score] and global participation coefficient (Pc).

| Dependent variables | Independent variables | *β* | 95% CI | *t* | *p* |
| --- | --- | --- | --- | --- | --- |
| Global Pc | FHD | 0.13 | [-0.17, 0.42] | 0.86 | 0.39 |
|  | Age | -0.06 | [-0.20, 0.09] | -0.78 | 0.44 |
|  | Sex | 0.02 | [-0.29, 0.34] | 0.16 | 0.87 |
|  | Years of education | 0.03 | [-0.12, 0.17] | 0.36 | 0.72 |
|  | Mean FD | 0.27 | [0.13, 0.42] | 3.68 | 0.0003 |
|  | no. of brain nodes | 0.32 | [0.18, 0.47] | 4.32 | < 0.0001 |
| Global Pc | CAIDE | -0.11 | [-0.26, 0.04] | -1.47 | 0.14 |
|  | Mean FD | 0.27 | [0.13, 0.42] | 3.73 | 0.0003 |
|  | no. of brain nodes | 0.30 | [0.15, 0.44] | 4.09 | < 0.0001 |

Standard coefficient β was reported. CI = confidence interval. FD = framewise displacement.
